# Understanding the difference in Diagnostic accuracy of MRI between ccRCC and pRCC :- A meta Analysis

**DOI:** 10.1101/2023.06.18.23291572

**Authors:** Dev Desai, Sanae Majdouli, Dwija Raval, Dev Andharia, Abhijay B. Shah, Hetvi Shah

## Abstract

**Background:** Renal cell carcinoma (RCC) comprises cancer originating from the renal epithelium and takes up for >90% of cancers in the kidney. The disease consists of >10 histological and molecular subtypes, of which clear cell RCC (ccRCC) is the most common and is responsible for most cancer-related deaths. In an attempt to ensure an early diagnosis to plan the further course of surgical intervention, pre-operative diagnosis plays an important role. MRI plays a crucial role in the diagnosis of RCC and planning for surgery for presumed RCC, especially for identifying enhanced soft tissue within renal lesions. The purpose of this meta-analysis is to ascertain the accuracy of MRI in diagnosing papillary renal cell carcinoma (pRCC) compared to clear cell carcinoma to reach a definitive diagnosis and thus, help in surgical intervention.

**Methods:** Medical literature was comprehensively searched and reviewed without restrictions to particular study designs, or publication dates using PubMed, Cochrane Library, and Google Scholar databases for all relevant literature. The extraction of necessary data proceeded after specific inclusion and exclusion criteria were applied. In this Meta-Analysis, a total of 5 papers involving 755 lesions were considered for Clear cell carcinoma. A total number of 13 papers regarding 1009 lesions of papillary renal cell carcinoma were considered. wherein two writers independently assessed the caliber of each study as well as the use of the Cochrane tool for bias risk apprehension. The statistical software packages RevMan (Review Manager, version 5.3), SPSS (Statistical Package for the Social Sciences, version 20), and Excel in Stata 14 were used to perform the statistical analyses.

**Results:** We calculated the sensitivity and specificity of MRI in diagnosing pRCC and ccRCC in the different papers, For the MRI in ccRCC, The sensitivity is 0.81 with a CI of 95% in a range of 0.77 to 0.86, the mean being 0.049. The Specificity of the MRI is 0.77 with a CI of 95% in a range of 0.68 to 0.86, the mean being 0.091. For the pRCC, The Sensitivity of the MRI in pRCC is 0.66 with a CI of 95% in a range of 0.52 to 0.80; the mean being (0.14). The Specificity of the MRI in pRCC is 0.87 with a CI of 95% in a range of 0.80 to 0.94, the mean being 0.072.

**Conclusion:** Magnetic resonance imaging (MRI) offers a thorough evaluation of renal masses that takes in to account both, functional factors and several types of tissue contrast. In light of the above mentioned clinical requirements, these characteristics of MRI have sped up the process of early detection, diagnosis, staging, and evaluation of the aggressiveness and therapeutic r esponse of RCC

## INTRODUCTION

Renal cell carcinoma (RCC) comprises cancer originating from the renal epithelium and takes up >90% of cancers in the kidney. The disease consists of >10 histological and molecular subtypes, of which clear cell RCC (ccRCC) is the most common and is responsible for most cancer-related deaths.[1]

10–15% of all renal cell carcinomas are Papillary renal cell carcinomas (PRCC), making it the second most common subtype after clear cell RCC.[2] In comparison to clear cell RCC, which is the most prevalent RCC subtype, papillary RCC (and in particular type 1 papillary RCC) is typically linked with lower histologic grade, a lower incidence of distant metastasis, and better cancer-specific survival. Clear cell RCC displayed a 5-year cancer-specific survival rate of 76.9% compared to papillary RCC’s 85.1%.[2] Despite various subtypes of renal cell carcinoma, the definitive management remains surgical resection of the tumor masses along with total or partial nephrectomy based on the severity of the tumor.

In an attempt to ensure an early diagnosis to plan the further course of surgical intervention, pre-operative diagnosis plays an important role. MRI plays a crucial role in the diagnosis of RCC and planning for surgery for presumed RCC, especially for identifying enhanced soft tissue within renal lesions.[2]Taking this into consideration, there are also a few limitations. Sensitivity and specificity reported in studies using the same MRI features to identify papillary RCC are highly variable.[2]Cancer can be diagnosed with a biopsy, but there is a significant non-diagnosis rate and risk of consequences.[3]It is important to note that there are some potential risks associated with a biopsy, including bleeding, infection, and damage to surrounding structures.

By analyzing the T2 image intensity, enhancement pattern, and apparent diffusion coefficient, MRI can assist distinguish papillary renal cell carcinoma from other neoplasms. Papillary RCC on MRI has been associated with signal loss on opposed-phase imaging, a substantial hyperenhancement on all dynamic contrast-enhanced MRI phases, and a hypointense T2 signal.[2]

Clear cell renal carcinoma typically appears as a well-defined mass with low signal intensity on T2-weighted images and high signal intensity on T1-weighted images. This is due to the presence of lipid and protein content within the tumor cells. In addition, areas of necrosis and hemorrhage within the tumor may also be visible on MRI.

However, it is important to note that the appearance of RCC on MRI can vary depending on several factors, including the stage and grade of the tumor, as well as the presence of other pathological features. Therefore, MRI is usually used in conjunction with other imaging modalities and clinical information to aid in the diagnosis and management of RCC.

The purpose of this meta-analysis is to ascertain the accuracy of MRI in diagnosing papillary renal cell carcinoma compared to clear cell carcinoma to reach a definitive diagnosis and thus, help in surgical intervention.

**(Figure 1).**
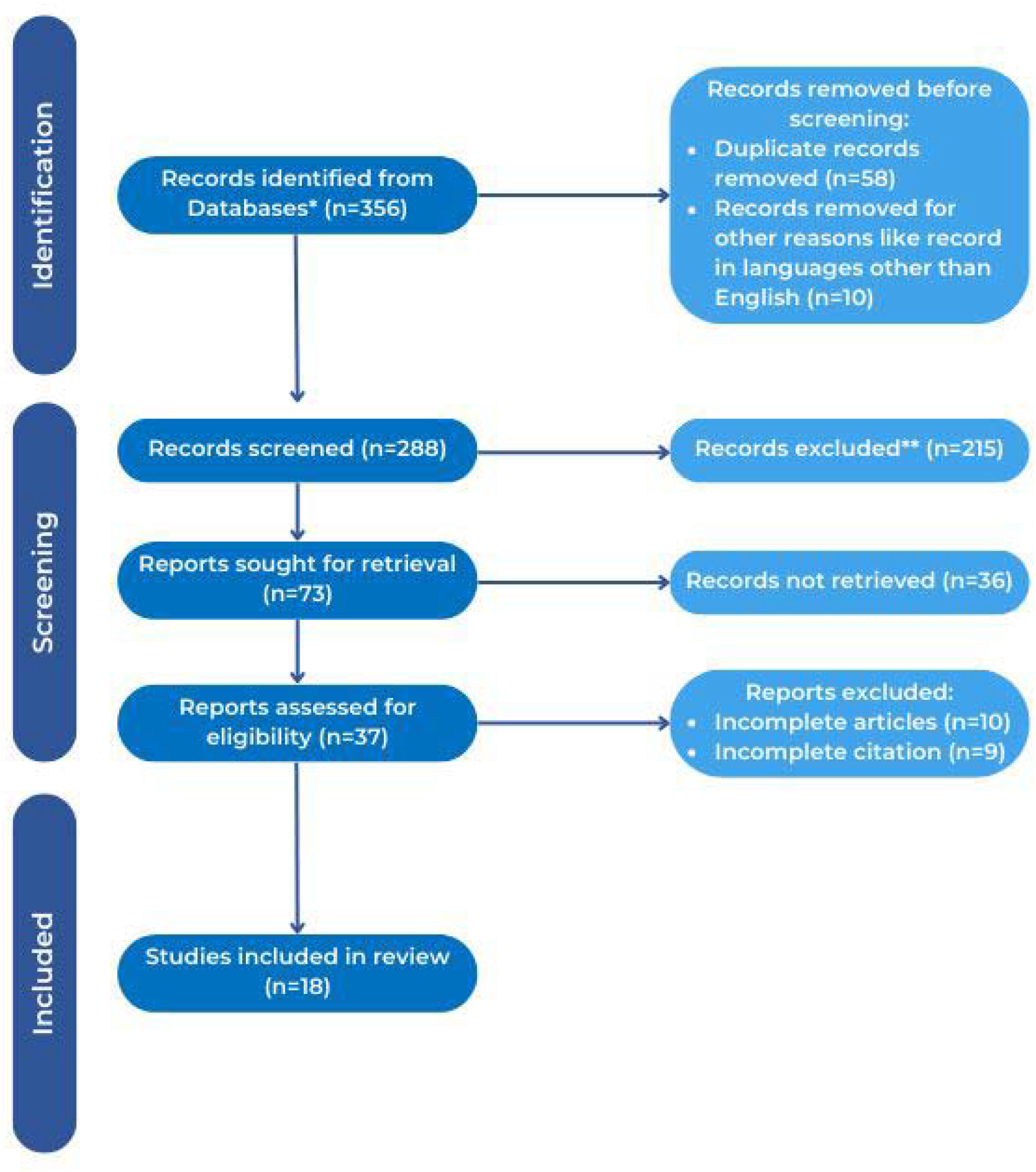
PRISMA Flowchart.

## METHODOLOGY

### DATA COLLECTION

To collect data, a search was done by two individuals on PubMed, Google Scholar, and Cochrane Library databases. Only Full-Text Articles written in English were considered. The medical subject headings (MeSH) and keywords “Renal cell carcinoma”, “Clear cell carcinoma”. “Papillary renal cell carcinoma”, “CT”, “MRI”, and “early detection” were used. References, reviews, and meta-analyses were scanned for additional articles.

### INCLUSION AND EXCLUSION CRITERIA

Titles and abstracts were screened, and duplicates and citations were removed. References of relevant papers were reviewed for possible additional articles (Snowballing). Papers with detailed patient information and statically supported results were selected.

We searched for papers tackling the diagnosis of clear cell carcinoma and papillary renal cell carcinoma using MRI as a diagnostic procedure.

The inclusion criteria were as follows:

1. Studies providing information about the accurate diagnosis of Renal cell carcinoma using MRI;
2. Studies tackling the diagnosis differentiation of Clear cell carcinoma and papillary renal cell carcinoma using MRI;
3. Studies published in English.

The exclusion criteria were:

1. Articles that were not full text;
2. Unpublished articles;
3. Articles in other languages than English.

### DATA EXTRACTION

Each included paper was independently evaluated by two reviewers. We extracted the number of patients, their age, procedure modality, and incidence of the predecided complications from each article. Further discussion or consultation with the author and a third party was used to resolve conflicts. The study’s quality was assessed using the modified Jadad score. A total of 5 papers involving 755 lesions were considered for Clear cell carcinoma. A total number of 13 papers regarding 1009 lesions of papillary renal cell carcinoma were considered, according to the Preferred Reporting Items for Systematic Reviews and Meta-Analyses (PRISMA).

### ASSESSMENT OF STUDY QUALITY

Using the QualSyst tool, two writers, independently, assessed the caliber of each included study. This test consists of 10 questions, each with a score between 0 and 2, with 20 being the maximum possible overall score. Two authors rated each article independently based on the above criteria. The interobserver agreement for study selection was determined using the weighted Cohen’s kappa (K) coefficient. For deciding the bias risk for RCTs, we also employed the Cochrane tool. No assumptions were made about any missing or unclear information.

### STATISTICAL ANALYSIS

The statistical software packages RevMan (Review Manager, version 5.3), SPSS (Statistical Package for the Social Sciences, version 20), Google Sheets, and Excel in Stata 14 were used to perform the statistical analyses. The data was obtained and entered into the analytic software. [4]Fixed-or random-effects models were used to estimate Sensitivity, Specificity, positive predictive value (PPV), diagnostic odds ratios (DOR), and relative risk (RR) with 95 percent confidence intervals to examine critical clinical outcomes (CIs). Diagnosis accuracy and Younden index were calculated for each result. Individual study sensitivity and specificity were plotted on Forest plots and in the receiver operating characteristic (ROC) curve. The forest plot and Fagan’s Nomogram were used to illustrate the sensitivity and specificity of different papers.

### BIAS STUDY

The risk of bias was evaluated using QUADAS-2 analysis. This tool includes 4 domains: Patient selection, Index test, Reference standard, Flow of the patients, and Timing of the Index tests.

## RESULT

### MRI in ccRCC

Table 1 presents all the descriptive data of the included papers. The forest chart (figure 2) summarizes a comparison of the sensitivity and specificity of MRI in diagnosing pRCC and ccRCC in the different papers. The comparison is depicted in the SROC curve (figure3). A total of 5 RCTs with 755 patients were selected for the study. The value of True positive is 363, True Negative is 241, False negative is 80, and False Positive is 71. With a confidence interval of 95%. Sensitivity, specificity, and Positive Predictive values were also calculated and summarized in Figure 2. The Sensitivity of the MRI is 0.81 with a CI of 95% in a range of 0.77 to 0.86, the mean being 0.049. The Specificity of the MRI is 0.77 with a CI of 95% in a range of 0.68 to 0.86, the mean being 0.091. The positive predictive value (PPV) for the MRI is 0.83 with a CI of 95% in a range of 0.77 to 0.89, the mean being 0.058.

**Table 1.**

| Author name | Year | Methodology | Total lesions | Papillary lesions | Clear cell carcinoma lesions | Sensitivity | specificity |
| --- | --- | --- | --- | --- | --- | --- | --- |
| Maryellen R. Sun [5] | 2009 | MRI vs. Pathology | 103 | 28 | 75 | 96 | 93 |
| F Cornelis [6] | 2014 | MRI vs. Pathology | 100 | 16 | 57 | 38 | 100 |
| Ivan Pedrosa [7] |  | MRI vs. Pathology | 79 | 15 | 48 | 80 | 94 |
| Hersh Chandarana [8] | 2012 | MRI vs. Pathology | 74 | 19 | 55 | 89 | 96 |
| Brindley David Cupido [9] | 2017 | MRI vs. Pathology | 50 | 17 | 20 | 100 | 100 |
| Kewale Sasiwimonphan [10] | 2012 | MRI vs. Pathology | 81 | 16 | 49 | 100 | 72.3 |
| M. Raquel Oliva [11] | 2009 | MRI vs. Pathology | 49 | 21 | 28 | 95.2 | 85.7 |
| Winnie Fu[12] | 2016 | MRI vs. Pathology | 544 | 87 | 425 | 91 | 93 |
| David D. Childs [13] | 2014 | MRI vs. Pathology | 166 | 50 | 102 | 48 | 87 |
| Kengo Yoshimitsu [14] | 2006 | MRI vs. Pathology | 66 | 9 | 57 | 89 | 70 |
| Catherine A. Murray [15] | 2016 | MRI vs. Pathology | 69 | 58 |  | 21 | 100 |
| Nicola Schieda [16] | 2016 | MRI vs. Pathology | 226 | 63 | 150 | 100 | 92.9 |
| Lothar Ponhold [17] | 2016 | MRI vs. Pathology | 34 | 8 | 21 | 88 | 85 |
| Jung Jae Park [18] | 2017 | MRI vs. Pathology | 236 |  | 168 | 89.7 | 88.2 |

**Figure 2.**
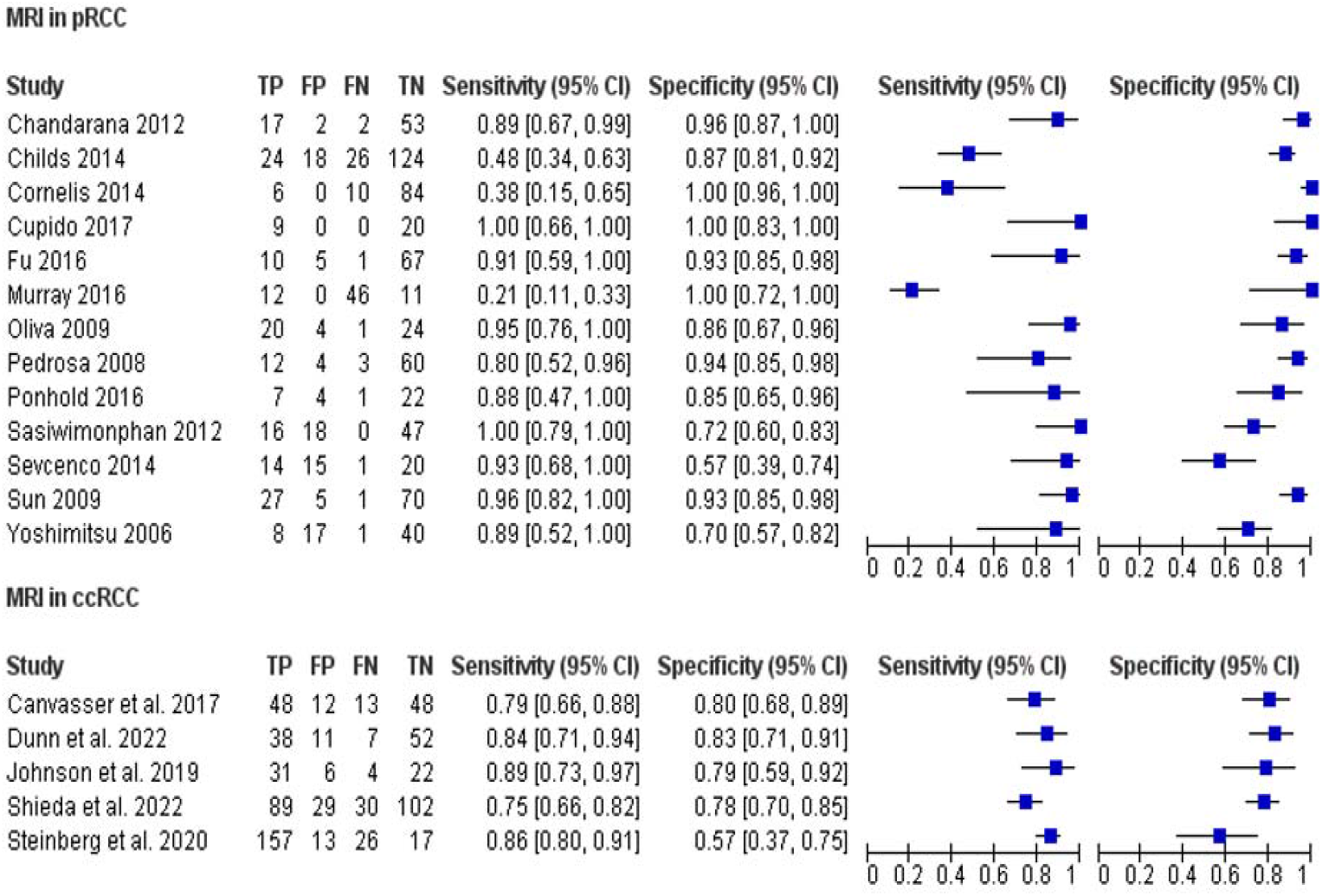
Forest plot for MRI in pRCC & ccRCC.

The summary of the ROC curve is illustrated in Figure 3. It shows that the area under the ROC (AUC) for MRI is 0.7959 and the overall diagnostic odds ratio (DOR) is 15.40. It also describes the Diagnostic Accuracy and The Younden index, which are 0.8 and 0.59 respectively.

**Figure 3.**
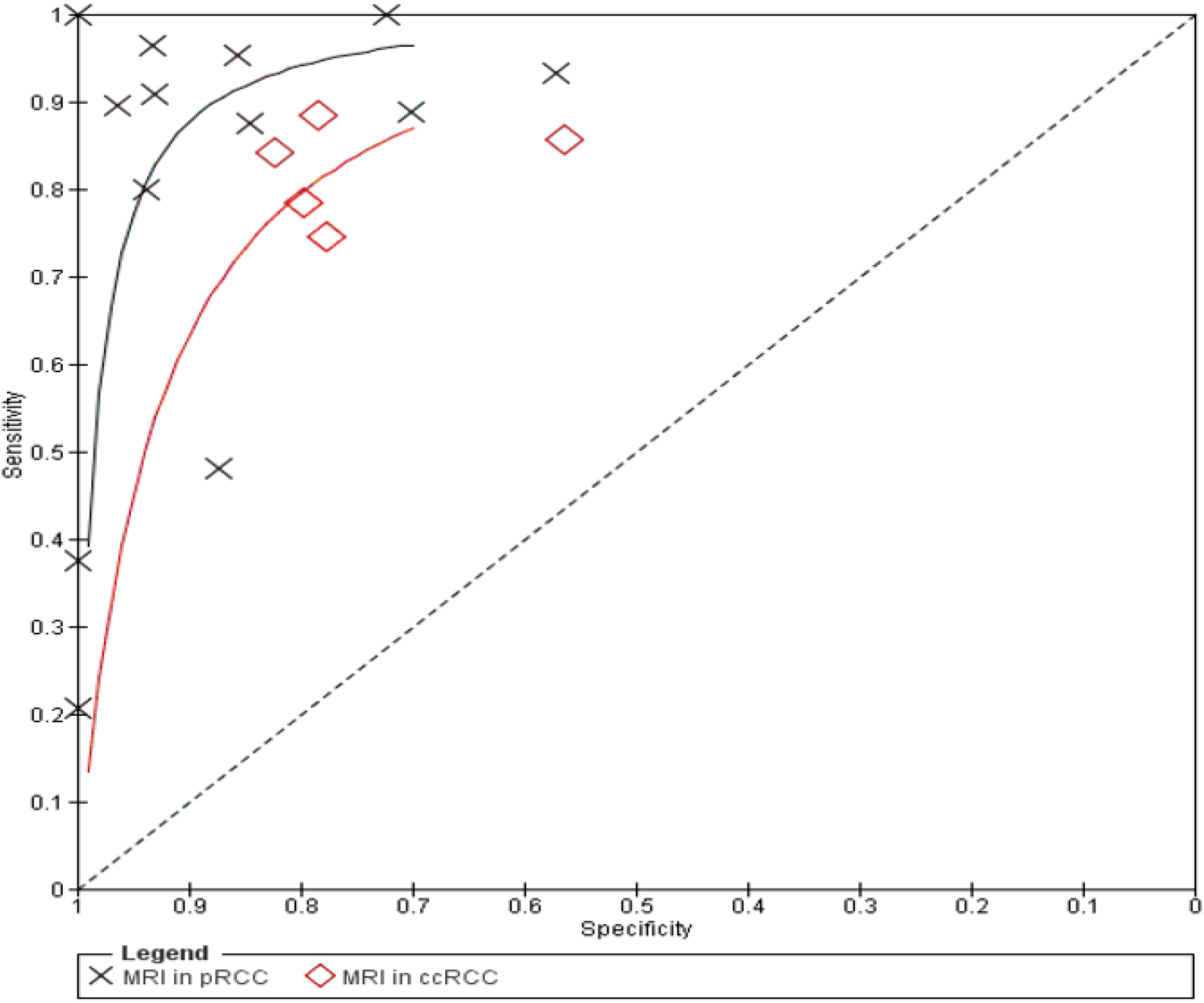
SROC curve for MRI in pRCC & ccRCC.

Figure 4 summarizes the Fagan plots analysis for all the included studies on MRI in ccRCC. It shows a Prior probability of 59% (1.4); a Positive likelihood ratio of 3.6; a probability of post-test 84% (5.1); a Negative likelihood ratio of 0.23, and a probability of post-test 25% (0.3).

**Figure 4.**
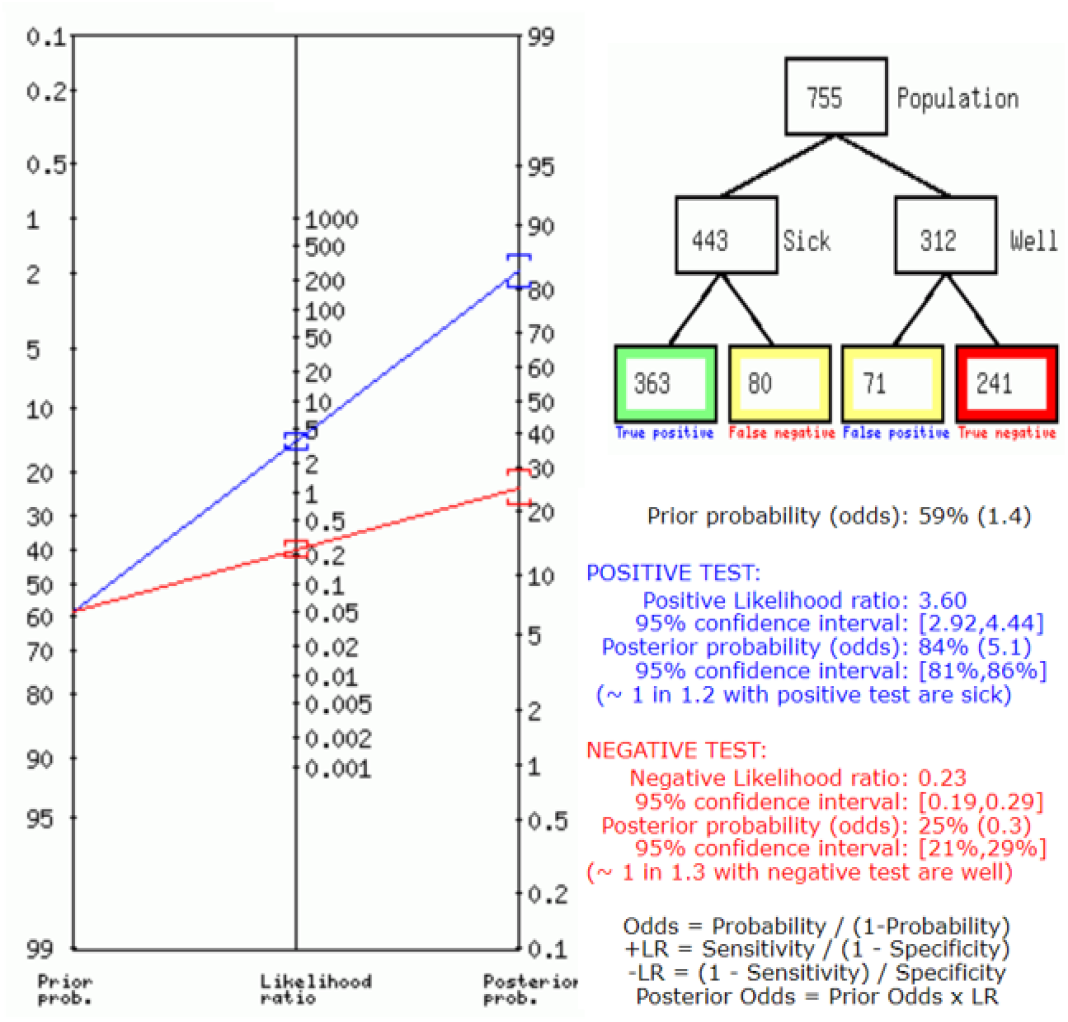
Fagan’s Analysis for ccRCC.

### MRI in pRCC

Above, Table 2 Describes all the description of papers used for the MRI in the pRCC studies. The forest chart (Figure 2) and the SROC curve (figure 3) present a comparison of the sensitivity and specificity of the included papers A total of 13 RCTs with 1009 patients were selected for the study. The value of True positive is 182, True Negative is 641, False Negative is 93, and False Positive is 93. With a confidence interval of 95%, Sensitivity, specificity, and Positive Predictive values were calculated and summarized in Figure 2. The Sensitivity of the MRI in pRCC is 0.66 with a CI of 95% in a range of 0.52 to 0.80; the mean being (0.14). The Specificity of the MRI in pRCC is 0.87 with a CI of 95% in a range of 0.80 to 0.94, the mean being 0.072. The positive predictive value (PPV) is 0.66 with a CI of 95% in a range of 0.54 to 0.78, the mean being 0.121.

**Table 2.** Bias Study

|  |  | Risk Of Bias |  |  |  | Applicability Concerns |  |  |
| --- | --- | --- | --- | --- | --- | --- | --- | --- |
| No.s | Author Name and Year | Patient Selection | Index Test | Reference Standard | Flow and Timing | Patient Selection | Index Test | Reference Standard |
| 1 | Sun | LOW | LOW | UNCLEAR | UNCLEAR | LOW | LOW | LOW |
| 2 | Cornelis | LOW | LOW | UNCLEAR | LOW | LOW | LOW | LOW |
| 3 | Pedrosa | LOW | LOW | UNCLEAR | LOW | LOW | LOW | LOW |
| 4 | Sevcenco | LOW | LOW | LOW | LOW | LOW | UNCLEAR | LOW |
| 5 | Chandarana | UNCLEAR | LOW | UNCLEAR | LOW | UNCLEAR | LOW | LOW |
| 6 | Cupido | UNCLEAR | LOW | HIGH | UNCLEAR | UNCLEAR | UNCLEAR | LOW |
| 7 | Sasiwimonphan | UNCLEAR | LOW | HIGH | UNCLEAR | UNCLEAR | UNCLEAR | LOW |
| 8 | Oliva | LOW | UNCLEAR | UNCLEAR | UNCLEAR | LOW | LOW | LOW |
| 9 | Fu | UNCLEAR | HIGH | HIGH | UNCLEAR | LOW | LOW | LOW |
| 10 | Childs | LOW | LOW | UNCLEAR | UNCLEAR | LOW | LOW | LOW |
| 11 | Yoshimitsu | UNCLEAR | UNCLEAR | UNCLEAR | UNCLEAR | LOW | LOW | LOW |
| 12 | Murray | LOW | LOW | UNCLEAR | UNCLEAR | LOW | LOW | LOW |
| 13 | Ponhold | UNCLEAR | LOW | LOW | UNCLEAR | LOW | UNCLEAR | LOW |

The summary of the ROC curve is described in Figure 3. It shows that the area under the ROC (AUC) for the MRI in pRCC is 0.7675 and the overall diagnostic odds ratio (DOR) is 13.52. Also, The Diagnostic Accuracy and Younden index are 0.81 and 0.53 respectively.

Figure 5 describes the summary of Fagan plots analysis for all the studies of pRCC. It shows a Prior probability of 27% (0.4), a Positive likelihood ratio of 5.22, a probability of post-test 66% (2.0), a Negative likelihood ratio of 0.39, and a probability of post-test 13% (0.1).

**Figure 5.**
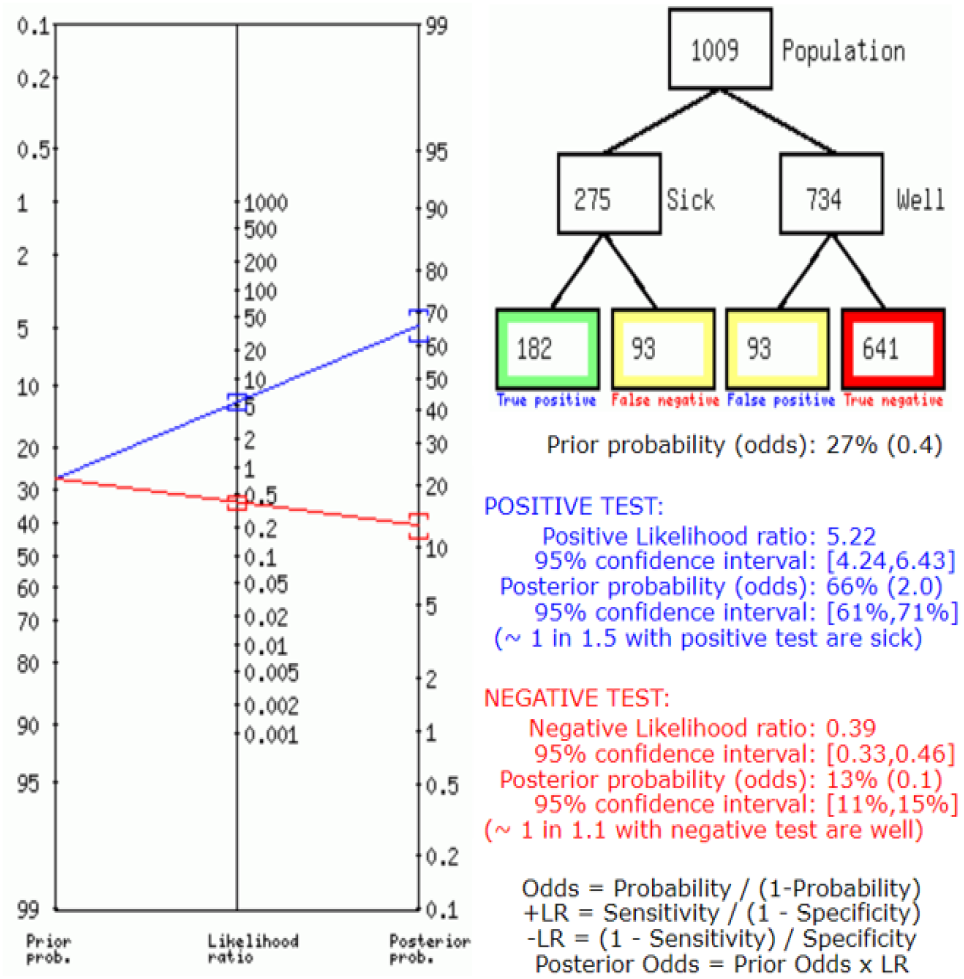
Fagan’s Analysis for pRCC.

### Bias Study

#### Publication Bias

The summary of publication bias for MRI in ccRCC and pRCC is shown in Table 2 and Figure 6. Patient selection bias was low in 7 studies and unclear in 6. The index test was low in 10 studies, unclear in 8, and high in 1. The reference standard was low in 2, unclear in 8, and high in 3. The flow and timing were low in 4 and unclear in 9. The applicability concerns in patient selection were low in 10 and unclear in 3. The index test was low in 9 and unclear in 4. The reference standard was low in 13.

**Figure 6.**
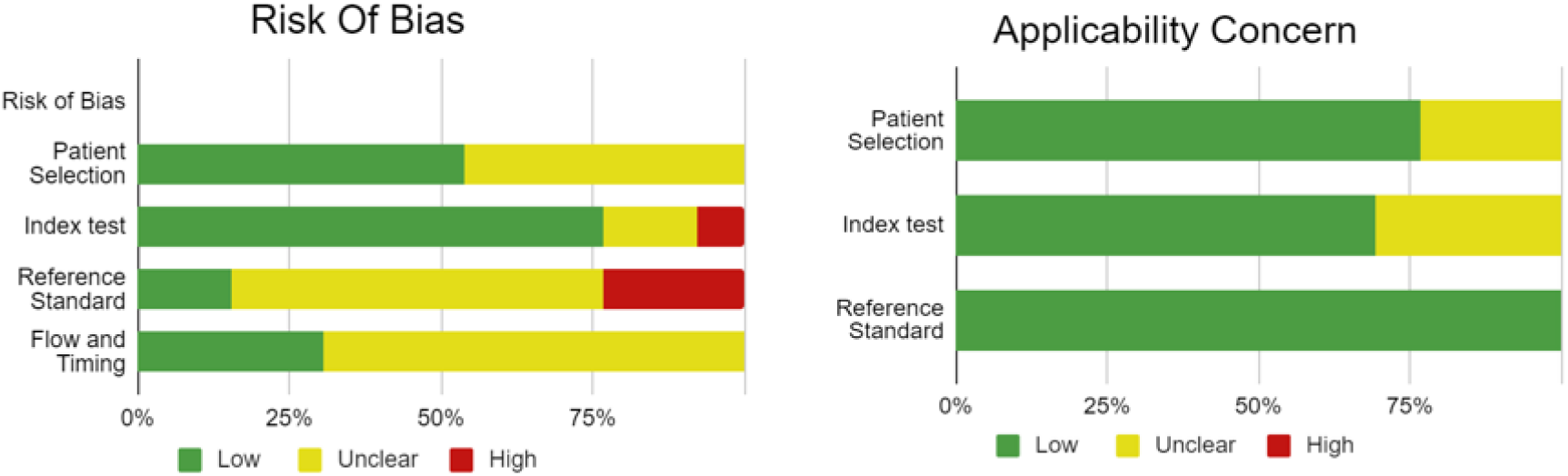
Summary of Bias Study.

## DISCUSSION

Renal cell carcinoma is a group of cancers originating from the renal epithelium and comprises >90% of renal cancers.[1]

Due to the common prevalence of renal cell carcinoma along with the diverse types and subtypes it encompasses, it becomes important to diagnose it accurately. However, this poses a great challenge for professionals considering the variations in histopathology, intensity, and malignant potential. Various methods are used to diagnose renal cell carcinoma, the major modality being imaging techniques, which include CT and MRI.

Many renal cell carcinomas are discovered incidentally when imaging is carried out for other disease conditions.

The biopsy is another diagnostic tool used. However, false negative results and other complications like bleeding and infection act as major challenges to taking this up as a primary tool for diagnosis. To deal with the challenges that percutaneous biopsy poses, MRI is used to rule out renal pathology.[19]

Magnetic resonance imaging (MRI) offers a thorough evaluation of renal masses that takes in to account both, functional factors and several types of tissue contrast. In light of the above-mentioned clinical requirements, these characteristics of MRI have sped up the process of early detection, diagnosis, staging, and evaluation of the aggressiveness and therapeutic response of RCC.The use of MRI in the treatment of both localized disease and advanced-stage disease has been made possible by technological advancements. We take up MRI procedures for imaging of renal masses, classifying them based on their types and subtypes as well as using it for the management of renal cell carcinoma.[20]

The sensitivity of MRI in diagnosing clear cell carcinoma is 81.9% and the specificity is 77.2%. In contrast to this, the sensitivity of papillary carcinoma is 66.2% while the specificity is 87.3%.

This demonstrates that MRI is a better tool to diagnose clear cell carcinoma. This plays an advantage because clear cell carcinoma is the most common type of renal cell carcinoma. Papillary carcinoma is the second most common type. [19]

The clear cell likelihood score (ccLS) is used for the diagnosis of clear cell carcinoma and has been found to have great accuracy. The scale helps in the diagnosis of both types, papillary and clear cell carcinoma. While the clear cell is diagnosed when ccLS score is more than 4, papillary renal cell carcinoma is diagnosed with a score greater than 1.[19] 10% to 15% of RCCs are pRCCs. Histologically, this subtype can be identified by its papillae, which have a fibrovascular core filled with foamy macrophages. Cystic degeneration, necrosis, and hemorrhage are frequently seen. Additionally, a well-formed tumor capsule is frequently visible.[20]

Despite clear cell carcinoma being the less aggressive of the two renal cell carcinomas, MRI has a relatively lower sensitivity in the diagnosis of this type. This is in contrast to clear cell carcinoma. This indicates the role of MRI in the diagnosis of both subtypes of renal cell carcinoma. This meta-analysis sought to address concerns regarding diagnostic modalities by determining the precision of MRI. This would allow for effective, efficient, and precise care of the condition and fulfill the need for effective action and avoid missed diagnoses in the clinical setting.

## Conclusion

Magnetic resonance imaging (MRI) offers a thorough evaluation of renal masses that takes in to account both, functional factors and several types of tissue contrast. In light of the above mentioned clinical requirements, these characteristics of MRI have sped up the process of early detection, diagnosis, staging, and evaluation of the aggressiveness and therapeutic response of RCC.

## Data Availability

All data produced in the present work are contained in the manuscript

